# Developing a prediction model for the risk of dissociative psychopathology from trauma and trait responsiveness to verbal suggestion

**DOI:** 10.64898/2026.05.11.26352886

**Authors:** Rebecca Morris, Madeline V. Stein, Lillian Wieder, Devin B. Terhune

## Abstract

**Background:** Dissociative experiences encompass a variety of discontinuities in awareness and perception that are elevated in the dissociative disorders and associated with extensive comorbid symptomatology. Accumulating evidence points to developmental trauma and trait responsiveness to verbal suggestions (REVS) as factors that confer risk for severe dissociative symptoms, but they have typically been studied in isolation. This study integrated these measures using prediction modelling to better understand their predictive value for the risk of dissociative psychopathology.

**Method:** 1,104 non-clinical participants completed measures of trauma, dissociation and trait REVS. The predictive model was developed using elastic net logistic regression, internally validated with 10-fold cross-validation, and assessed using receiver operating characteristic (ROC) curve and area under the ROC (AUROC). Variables entered into the model were components of REVS, trauma, age, and their interactions.

**Results:** A dissociative psychopathology at-risk group (7%) was characterised by younger age, greater trauma and elevated REVS, particularly involuntariness during cognitive-perceptual suggestions. The prediction model retained nine of ten predictors, with an AUROC of .77 [95% CI: .73, .82], reflecting good discrimination with moderate sensitivity (78%) but modest specificity (67%).

**Conclusions:** These findings reinforce trauma and trait REVS as risk factors for dissociative psychopathology and demonstrate that they can be integrated in a model that can identify at-risk individuals. Further validation and extension of the model is necessary to improve the identification of individuals at risk for severe dissociative symptomatology and the diagnosis of dissociative disorders with implications for outcome trajectories.

## 1. Introduction

Dissociation encompasses a constellation of symptoms involving a discontinuity of experience, with disturbances spanning dimensions of consciousness, identity, and memory (Sar, 2011; Spiegel et al., 2011). Although the dissociative disorders (DDs) are a discrete diagnostic category, they are transdiagnostic symptoms present in a diverse array of psychiatric conditions with extensive comorbidities (Chien & Fung, 2022; Langeland et al., 2020; Lyssenko et al., 2018; Sar, 2011). The complexity of symptom presentation, combined with limited awareness in clinical research and practice, results in frequent under- and mis-diagnosis, and significant delays until diagnosis (Langeland et al., 2020; Reinders & Veltman, 2020). These factors contribute to excessive patient suffering, implementation of inadequate treatment, and increased risk of comorbidities and suicidal behaviour (Bruno et al., 2025; Gleaves & Reisinger, 2023; Lundin, 2024; Shakya et al., 2024).

There is broad consensus that exposure to developmental trauma is among the most salient antecedents of dissociative psychopathology (McGuinness et al., 2025; Vonderlin et al., 2018). The trauma model (TM) of dissociation posits that dissociative symptoms serve as an internal coping mechanism in response to developmental trauma (Dalenberg et al., 2012; Gershuny & Thayer, 1999; Nijenhuis et al., 1998; Putnam, 1997). In particular, dissociative symptoms spanning depersonalisation to identity disturbances are hypothesised to alleviate the emotional effect of trauma with protective altered states of consciousness (Boysen, 2024; Buchnik-Daniely et al., 2021; Loewenstein, 2018). Empirical research, including neurobiological evidence, supports a role for trauma in the emergence of dissociative symptoms across clinical and non-clinical samples (Chalavi et al., 2015; Kate et al., 2020; King et al., 2020; Kira et al., 2022), with a more recent study highlighting the predictive value of trauma in the experience of dissociative symptoms (McGuinness et al., 2025).

Although neglected by the trauma model, trait responsiveness to verbal suggestion (REVS) has long figured prominently in models of dissociative psychopathology (Butler et al., 1996; Dell, 2017; Wieder et al., 2023). Suggestions refer to communications for a change in experience that is often framed as something that will happen to an individual (Kirsch, 1999; Stein et al., In Preparation). Elevated REVS (also known as direct verbal suggestibility (Oakley et al., 2021)) is hypothesised to predispose individuals to dissociative symptoms and disorders (Bell et al., 2011; Butler et al., 1996; Dell, 2017) by conferring risk for posttraumatic stress symptoms, or a broader proneness to the perturbation of awareness (Dell, 2021). In support of this hypothesised link, REVS has been shown to reliably correlate with posttraumatic stress symptoms (Keuroghlian et al., 2010; Sapkota et al., 2020; Yard et al., 2008) and meta-analyses have presented evidence for elevated REVS in dissociative disorders, trauma and stressor related disorders, and functional neurological disorder (Wieder et al., 2021; Wieder et al., 2022). REVS has specifically been flagged as a valuable method for differential diagnosis of dissociative disorders and psychosis (Mertens & Vermetten, 2018).

An outstanding question is whether and how developmental trauma and REVS interact in the emergence of dissociative symptoms. REVS, as measured by standardised scales involving the administration of direct verbal suggestions, does not reliably correlate with self-reported trauma (Nash et al., 1993; Putnam et al., 1995; Sapkota et al., 2020; Wieder & Terhune, 2019). One interpretation of the interaction of these variables is provided by the Diatheses Stress model (Butler et al., 1996), which posits that hypnotic suggestibility, a form of REVS, *moderates* the extent to which trauma elicits a dissociative response. In support of this model, multiple studies have found evidence that trauma and REVS interact in the prediction of dissociation, which is consistent with the proposal that higher REVS confers greater risk for dissociative psychopathology in response to trauma (Putnam et al., 1995; Wieder & Terhune, 2019).

Considering the negative outcome trajectories in the DDs, it is necessary to develop prediction models (Salazar de Pablo et al., 2021) that reliably identify patients who are at-risk for dissociative psychopathology (Srinivasan et al., 2022). Research using prediction modelling in dissociation is sparse, with most research focusing on dissociation as a predictor of other symptoms (Greene, 2018; Guzman Torres et al., 2023; Srinivasan et al., 2022). Nevertheless, preliminary research has replicated the finding that trauma can reliably predict dissociation (Kate et al., 2021; King et al., 2020; McGuinness et al., 2025). In particular, a recent machine-learning study moved beyond the narrow focus on trauma and found that childhood trauma, mental health symptoms (anxiety and depression) and other factors could be used to collectively predict dissociation with evidence for differential efficacy of predictors across age groups (McGuinness et al., 2025).

The present study sought to build upon prediction models of dissociative psychopathology by incorporating measures of trauma and REVS using elastic net logistic regression (Zou & Hastie, 2005) and receiver operating characteristic (ROC) curve analysis. We expanded upon the methods of previous studies by using an automated measure of REVS (*Brief Suggestibility Scale* [BSS]; (Wieder & Terhune, 2019)) that ensures standardisation in the administration of suggestions and circumvents experimenter effects (Acunzo & Terhune, 2021). The BSS assesses both behavioural responses and involuntariness during REVS, which is salient given that an attenuation in the sense of agency is a cardinal feature of dissociation ((Dell, 2010); see also (Brown et al., 2008)). In recognition of the complex factor structure of REVS, which includes a core ability and multiple subordinate abilities corresponding to specific suggestions (e.g., hallucinations (Barnier et al., 2022; Woody et al., 2005; Zahedi & Sommer, 2022)), we fit bifactor models (Bornovalova et al., 2020; Watkins, 2018) to BSS scores. This is important given that individuals with dissociative disorders tend to display greater REVS for dissociative and germane symptoms (e.g., amnesia; (Bryant et al., 2001; Wieder et al., 2021) and dissociation is reliably associated with hallucination-proneness (Bloomfield et al., 2021; Pilton et al., 2015). Our broad predictions were that trauma and REVS would independently and interactively predict risk for dissociative psychopathology and certain dimensions of REVS including involuntariness and response to hallucination suggestions would have greater predictive utility; these predictions were not pre-registered.

## 3. Methods

### 3.1. Participants

Participants (*N* = 1104) included non-clinical individuals (female: 669 [61%]; male: 407 [37%]; other: 19 [2%], missing: 9 [1%]; *M*_age_=35.35, *SD*=11.80 [*n*=1 missing]). The sample was drawn from four previous studies varying in size from 208 to 300 (Stein, Holt, et al., 2023; Stein, Wieder, et al., 2023; Stein, Wieder, et al., 2025; Wieder & Terhune, 2019). All participants provided informed consent in accordance with local ethical approval.

### 3.2 Measures

#### Trauma

The *Trauma Experiences Checklist* (TEC; (Nijenhuis et al., 2002)) is a 29-item self-report questionnaire used to measure exposure to traumatic events, such as sexual, emotional, and physical abuse. Participants responded in a binary fashion, with a total score ranging from 0 to 29. One dataset (*n* = 299 [27.08%]) items 28 and 29 of the TEC. Thus, TEC scores were transformed to percentages to allow comparability across the four datasets. Evidence substantiates the reliability and validity of the TEC (Nijenhuis et al., 2002) and it demonstrated good internal consistency in this sample (IZ = .79).

#### Dissociation

The *Dissociative Experiences Scale* (DES; (Carlson & Putnam, 1993)) is a 28-item self-report measure designed to assess dissociative experiences. Participants rate each item from 0-100%, in 10% increments, from “never” to “always”. The DES includes subscales indexing amnesia, absorption, and depersonalisation-derealisation. The DES has been validated across clinical and non-clinical samples, with high convergent and predictive validity (Carlson & Putnam, 1993; Vanijzendoorn & Schuengel, 1996), for a review, see (Wainipitapong et al., 2025), and displayed strong internal consistency in this sample (total: IZ = .94; amnesia: IZ = .81; absorption: IZ = .85; depersonalisation: IZ = .85).

#### REVS

The *Brief Suggestibility Scale* (BSS; (Wieder & Terhune, 2019)) is a computerised behavioural scale measuring non-hypnotic REVS or direct verbal suggestibility (Oakley et al., 2021). The BSS includes including six verbal suggestions for change in ideomotor responses (e.g., arm heaviness), motor control (e.g., arm paralysis), and perception (e.g., auditory hallucination). Participants rate the extent to which they behaviourally responded to each suggestion using a visual analogue scale that is subsequently scaled from 0 to 1, followed by a 6-point Likert scale to rate their perceived involuntariness (0 = did not experience at all, 1 = voluntary, 5 = involuntary-automatic) in order to capture the classic suggestion effect (Bowers et al., 1988; Wieder & Terhune, 2019). Behavioural and scales are integrated by computing the mean of the *z*-transformed scores to form a composite score (BSS-C; Wieder & Terhune, 2019). The BSS displayed good internal consistency in the current sample (behavioural: IZ = .77; involuntariness: IZ = .78; BSS-C: IZ = .79).

### 3.3 Procedure and Ethics

The study combines data from four previous cross-sectional studies, leading to slight variations across procedures. Participants were recruited through Prolific, a web-based participant recruitment platform (www.prolific.com), reviewed an information sheet, and provided informed consent. Participants provided demographic information and then completed the measures in counterbalanced order via Qualtrics (www.qualtrics.com). Participants were debriefed and compensated between £3.35- £5/hour (Stein, Holt, et al., 2023; Wieder & Terhune, 2019).

### 3.4 Statistical Analyses

The data is publicly available here: https://osf.io/sa6bw. All analyses were performed in *R* (v. 4.5.1 ; (R Core Team, 2023), IBM SPSS (v. 31; (IBM Corp, 2025), and Mplus (Muthén & Muthén, 1998–2017). Trauma scores were transformed to percentages to address the missing data for items 28 and 29 on the TEC in one of the datasets used (*n* = 299 [27.08%]). Univariate outliers were winsorised at the 90^th^ percentile to minimise the impact of extreme values; multivariate outliers (Mahalanobis distance, *p*<.001) were identified, but not removed or adjusted, due to the large proportion (*n* = 101 [9.15%]) and the potential to remove participants at the boundaries of dissociation.

We first sought to discern the latent factor structure of the BSS by fitting exploratory correlated factors and bifactor models to its behavioural and involuntariness items.

Correlated factor analysis identifies non-orthogonal latent variables that explain variance in observed variables (Watkins, 2018), whereas bifactor analysis identifies a general (*g*) factor that reflects a core latent variable that explains variance in all or most items, alongside specific factors representing subsets of items (Bornovalova et al., 2020). The final model was selected based on parsimony and fit indices: Tucker Lewis Index and Comparative Fit Index >.90, Root Mean Square Error of Approximation <.05, and Standardises Root Mean Square Residual <.08.

The outcome measure in the prediction model (risk for dissociative psychopathology) was quantified by computing the probability of membership in the DES taxon (DES-T), a subset of items previously linked to severe dissociative symptomatology (Waller et al., 1996) and then binarising this probability value using a conservative *p*(taxon) threshold of .95.

Elastic net logistic regression was then performed to evaluate the prediction model and included REVS measures (BSS-C, BSS behavioural general factor [BSS:B-G], BSS involuntariness general factor [BSS:I-G], BSS involuntariness perceptual factor [BSS:I-PF]), TEC, and age. In addition, interaction terms were created using the product of *z*-scores of REVS and trauma (BSS-C×TEC; BSS:I-PF×TEC), age and trauma (AGE×TEC), and age and REVS (AGE×BSS-C).

Elastic net (Zou & Hastie, 2005) combines ridge (α = 0) and LASSO (α = 1) penalties, where alpha determines the combined weight and lambda determines the strength of regularization, to identify variables that maximise prediction accuracy, whilst excluding those with zero coefficients for a parsimonious solution. The model was internally validated with 10-fold cross-validation, where the dataset is apportioned into 10 folds (*n* = 110), training the model on nine and validating on the remaining fold, and iterated ten times. Model discrimination was visualised using receiver operating characteristic (ROC) curve and quantified by the area under the ROC (AUROC), ranging from .5 to 1, corresponding to chance level to perfect discrimination, respectively. Benchmarks proposed by Hosmer Jr et al. (2013) include: .70-.79 (acceptable); .80-.89 (excellent); and .90-1 (outstanding).

Youden’s Index (Youden, 1950) determines the optimal cut-off to balance sensitivity (correct classification of risk for dissociative psychopathology), and specificity (correct classification of non-risk for dissociative psychopathology). Overall model performance was evaluated using the Brier score (range: 0-0.25 for a perfect to non-informative model) and the scaled Brier score (range: 0-100% for non-informative to a perfect model) (Steyerberg et al., 2010).

## 4. Results

### 4.1. Latent factor structure of REVS

We sought to characterise the latent structure of REVS by fitting multiple bifactor models to the BSS behavioural and involuntariness subscales (see Supplementary Materials). The analyses indicated that a single general factor model provided the best fit for the behavioural subscale (BSS:B-G), whereas a two-factor solution provided the best fit for the involuntariness subscale, comprising a general factor (BSS:I-G) and a perceptual suggestion factor (BSS:I-PF).

### 4.2. Sample characteristics

7% (*n* = 77) of the sample were classified as at-risk for dissociative psychopathology (*p*(taxon) ≥ 0.95). As can be seen in Table 1, the at-risk group were significantly younger, reported greater trauma, and displayed greater scores on all BSS and DES measures but the two groups did not significantly differ in gender distributions.

**Table 1.**
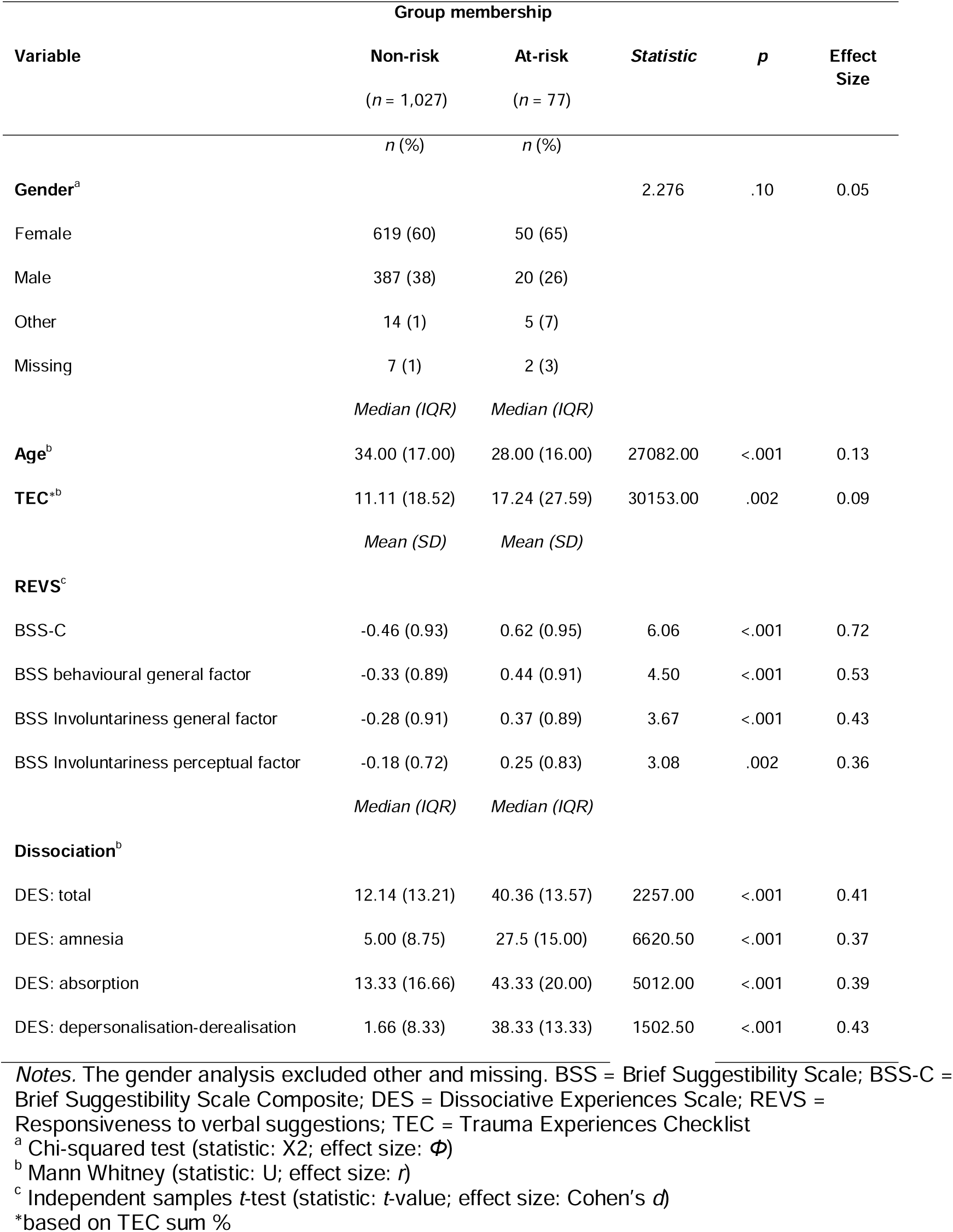
Demographic and psychological measures as a function of dissociative psychopathology risk membership (N=1,104).

### 4.3. Prediction Modelling

A prediction model for dissociative psychopathology risk using REVS measures, trauma, age, and their interaction terms was evaluated. Elastic net logistic regression with 10-fold cross-validation identified the optimal alpha (α = .5) and lambda (λ = .006), retaining nine of the ten predictors with only age reaching significance (see Table 2). The ROC curve yielded an AUC of .77 [95% CI: .73, .82], indicating a 77% probability that the model correctly classifies individuals with regard to their risk for dissociative psychopathology (see Figure 1). The optimal cutoff, determined by the Youden Index (*J* = .45, *SE* = .05), indicates good discriminatory ability of the model, with good sensitivity (78%) but modest specificity (67%). The brier score of .06 reflects excellent model fit, and the scaled brier score indicates that the predictor set explained 75% of the mean squared error between predicted and observed outcomes.

**Figure 1.**
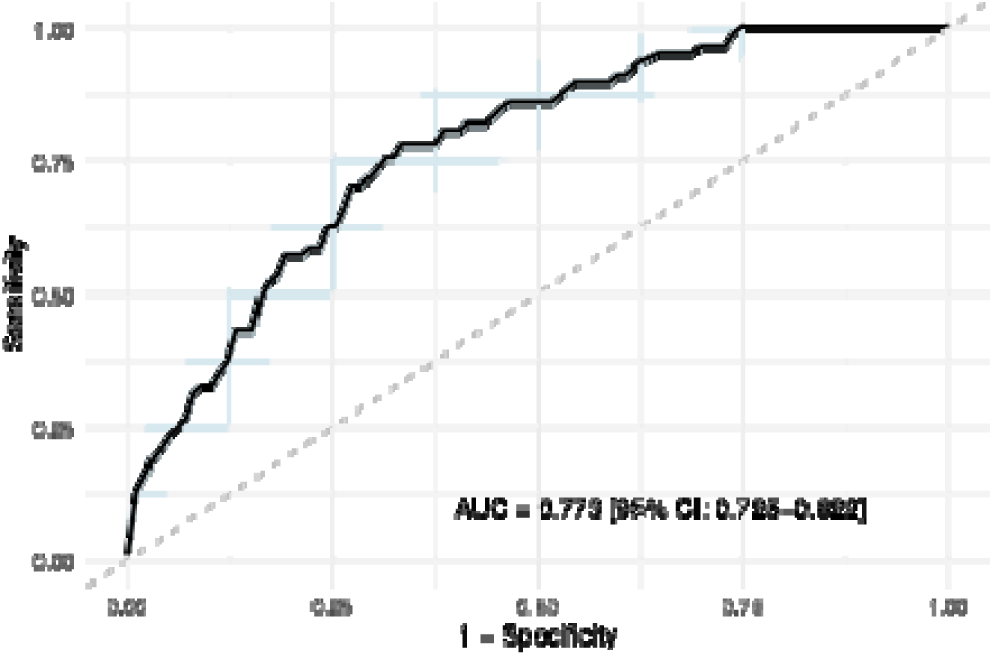
Receiver operating characteristic (ROC) curve of the prediction model of risk for dissociative psychopathology (N = 1,104). *Notes.* Receiver operating characteristic (ROC) curve for the prediction model plots the true positive rate (sensitivity) against the false positive rate (1- specificity). The solid diagonal line represents random chance level discrimination. Confidence intervals (CIs) are reflected by the light blue shading.

**Table 2.**
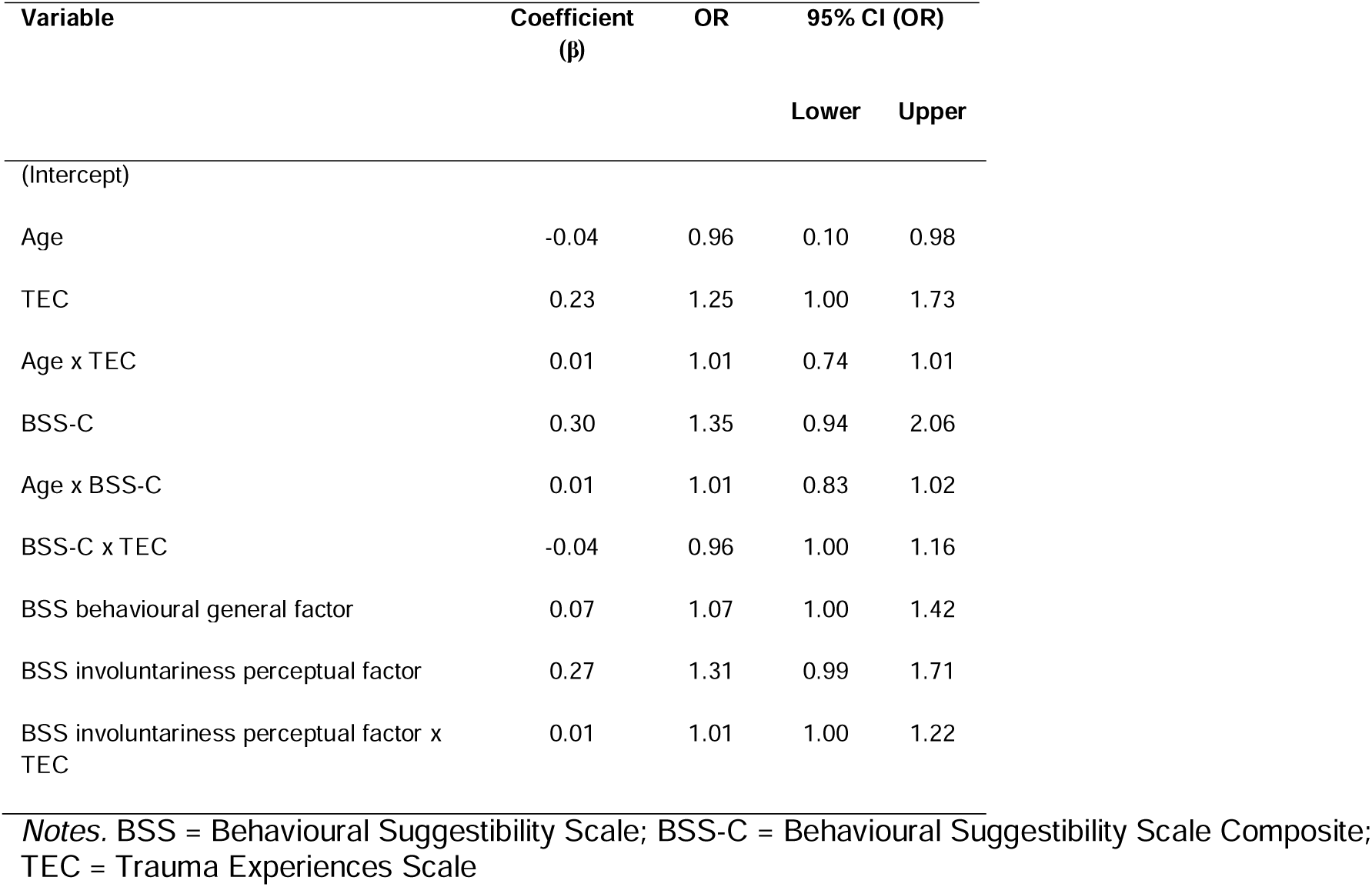
Elastic Net Logistic Regression Model Coefficients and Odds Ratios with 95% CI for the retained predictor variables in the prediction of risk for dissociative psychopathology (N=1,104).

| Variable | Coefficient<br>($\beta$) | OR | 95% CI (OR) | |
| --- | --- | --- | --- | --- |
|  |  |  | Lower | Upper |
| (Intercept) |  |  |  |  |
| Age | -0.04 | 0.96 | 0.10 | 0.98 |
| TEC | 0.23 | 1.25 | 1.00 | 1.73 |
| Age x TEC | 0.01 | 1.01 | 0.74 | 1.01 |
| BSS-C | 0.30 | 1.35 | 0.94 | 2.06 |
| Age x BSS-C | 0.01 | 1.01 | 0.83 | 1.02 |
| BSS-C x TEC | -0.04 | 0.96 | 1.00 | 1.16 |
| BSS behavioural general factor | 0.07 | 1.07 | 1.00 | 1.42 |
| BSS involuntariness perceptual factor | 0.27 | 1.31 | 0.99 | 1.71 |
| BSS involuntariness perceptual factor x TEC | 0.01 | 1.01 | 1.00 | 1.22 |

Notes. BSS = Behavioural Suggestibility Scale; BSS-C = Behavioural Suggestibility Scale Composite; TEC = Trauma Experiences Scale

Our final analysis sought to corroborate the model using an alternative classification of dissociative psychopathology risk (DES>30; (Carlson et al., 1993; Lyssenko et al., 2018). The binarized DES-T and DES-30 threshold scores were highly and significantly correlated (φ = .608, *p*<.001), suggesting that the two indices capture a very similar underlying phenomenon. The Elastic net regression model using binarized DES-30 threshold scores as the outcome yielded nearly identical results to the main model (AUC = .762 [95% CI: .722, .802]; see Supplementary Materials). BSS-C and age were the only significant predictors among those retained in the model. Taken together, these results suggest that risk for dissociative psychopathology can be predicted to a moderate level using a multivariate model including age, trauma, and REVS and that these results are not dependent upon a specific classification of dissociative psychopathology.

## 5. Discussion

This study sought to develop a multivariate model to predict risk for dissociative psychopathology in a large non-clinical sample. The results demonstrated that a model incorporating trauma, age, REVS, and their interactions, can predict risk for dissociative psychopathology with 77% classification accuracy. These results align with research demonstrating reliable links between REVS and various forms of dissociative psychopathology (Wieder et al., 2022) and reaffirms the diagnostic value of REVS when identifying DDs (Mertens & Vermetten, 2018). Alongside other prediction models incorporating stress and mental health symptoms (McGuinness et al., 2025), this work may help to lay the foundation for the reliable identification of individuals at risk for dissociative psychopathology.

Using a conservative threshold derived from the probability of DES taxon membership (DES-T; (Waller et al., 1996)), we observed a prevalence rate of 7% for risk of dissociative psychopathology. This risk rate aligns with the 7% estimate for dissociative psychopathology among college populations (Kate et al., 2020). Given that this is an estimate of risk rather than pathology *per se*, it is broadly congruent with the estimated prevalence of dissociative disorders in the general population (1-2%; (Boyer et al., 2022; Şar et al., 2007; Yang et al., 2023)) as not all at-risk individuals will meet formal diagnostic criteria.

Our results firstly demonstrate that individuals at-risk for dissociative psychopathology have a unique trauma and cognitive profile relative to those who are not at risk. Participants classified as at-risk reported significantly higher levels of traumatic experiences, as measured by the TEC, consistent with prior evidence (Kluemper & Dalenberg, 2014; Luoni et al., 2018; Rafiq et al., 2018). Moreover, at-risk individuals demonstrated significantly heightened REVS, in line with prior data (Dell, 2017; Wieder et al., 2023). In direct comparisons of the at-risk and not-at-risk groups, BSS-C scores displayed the largest effect size, which was notably higher than the trauma measure. Importantly, the effect size for the BSS-C was larger than that for the BSS behavioural general factor, plausibly reflecting the added benefit of incorporating an assessment of involuntariness in the measurement of REVS. This result is salient given that aberrant sense of agency is commonly observed in dissociative psychopathology (Dell, 2010). Moreover, this aligns with previous results demonstrating that individuals who are both high in dissociation and REVS exhibit greater involuntariness when responding to suggestions (Terhune et al., 2011). These findings support the idea that dissociative psychopathology may reflect a constellation of trauma-related and cognitive characteristics that differentiate at-risk individuals from those not at-risk and warrants moving away from simple trauma models to multivariate models incorporating cognitive risk factors, trauma exposure, and relevant demographic characteristics (see also (McGuinness et al., 2025).

The prediction model retained nine variables of those in the predictor set. All predictors except age, were not significant, which is likely due to high multicollinearity between variables and the application of the shrinkage factor within the elastic net regression model. The retention of trauma in the model aligns with findings that trauma exposure significantly predicts dissociation (Kate et al., 2020; McGuinness et al., 2025; Vonderlin et al., 2018), documenting the predictive value of trauma in risk for dissociative psychopathology, which may guide future evaluations for the pathophysiology of DDs. However, trauma was a weak predictor, despite being the only trauma variable included in the model, indicating instability and a lack of precision in the estimated predictive value of trauma, aligning with evidence indicating that trauma is insufficient to explain most of the variance in dissociation (Boysen, 2024; Briere et al., 2005; McGuinness et al., 2025). On the other hand, this observed imprecision is potentially attributable to our use of the TEC, which indexes a variety of different forms of trauma, and thereby may be less sensitive than measures targeting childhood trauma, which has been shown to be a more robust predictor of dissociation (McGuinness et al., 2025; Vonderlin et al., 2018). Further, one dataset in our analysis was missing two TEC items, which pertained to forms of sexual abuse. Whilst other items also capture forms of sexual abuse, participants missing this data may have an underestimated traumatic exposure. Such underestimation would reduce variability in trauma scores, contributing to the observed weak predictor. Prior research documents individuals with DD who have not experienced trauma (Merckelbach & Patihis, 2018), suggesting that trauma may only be relevant in extreme cases of dissociative psychopathology (Dalenberg et al., 2012). Insofar as we used a conservative DES-T cutoff of .95, the current study does not discriminate between severity of the at-risk cases, and therefore, cannot be used to determine whether trauma is only relevant in extreme cases of dissociative psychopathology.

All measures of REVS, except for the BSS general involuntariness factor, were retained in the model. This aligns with meta-analytic research demonstrating elevated REVS in DDs and germane conditions than non-clinical controls (Wieder et al., 2023; Wieder et al., 2022). These results are at odds with claims that dissociation is unrelated to suggestibility (Brand et al., 2016; Lynn et al., 2012), but do not corroborate the view that dissociative psychopathology is driven by iatrogenesis and false memory formation (Lynn et al., 2014; Lynn et al., 2019). The latter phenomena are most closely tied to interrogative suggestibility, which is distinct from REVS (Polczyk, 2016; Stein, Faerman, et al., 2025) and not atypical in the DDs (Vissia et al., 2016). We maintain that the tendency to view REVS and its putative link to dissociative psychopathology through the narrow lens of iatrogenesis and false memory has hindered theoretical progress in the study of dissociative psychopathology (Wieder et al., 2023).

Rather, the results suggest that individuals who have liminal states of awareness and perception that are reliably modulated through verbal suggestions seem to be at greater risk for dissociative psychopathology (Butler et al., 1996) or that proneness to dissociative experiences confers greater REVS (Wieder et al., 2023; see also (Dell, 2021)). In line with the former (diathesis stress) model (Butler et al., 1996), interaction effects between trauma and BSS-C, and trauma and the BSS involuntariness perceptual factor, were retained in the model although these were non-significant, weak effects. This broadly aligns with preliminary findings showing the interaction of trauma and REVS to predict dissociation (Wieder & Terhune, 2019).

REVS may vary as a function of dissociation severity or constellations of dissociative phenomena (Brown, 2006; Holmes et al., 2005). By using a single cut-off based on the DES-T, this study was unable to discriminate between severity and symptom constellations. It has been theorised that REVS mechanistically aligns more with compartmentalisation (e.g., identity disturbances) than detachment (e.g., depersonalisation) dissociative symptoms (Brown, 2006). Indirect evidence in support of this hypothesis comes from a recent study that observed typical REVS in patients with depersonalisation-derealisation disorder, given that these individuals are primarily characterised by detachment symptoms (Millman et al., 2022). Moreover, evidence for elevated REVS in dissociative psychopathology primarily comes from functional neurological disorder and dissociative identity disorder, which are characterised by compartmentalisation phenomena (Wieder et al., 2022). The DES-T primarily comprises compartmentalisation symptoms, but not entirely, and thus our results are somewhat equivocal regarding this distinction. Thus, REVS likely varies across dissociation symptom constellations, which may be obscured when using broad at-risk categorisation based on the DES-T.

All three age variables were retained in the model, with the main effect of age highlighting that dissociative experiences are lower with increasing age (Conner, 2015; Torem et al., 1992; Walker et al., 1996). The retention of the age interaction terms suggests a moderating role of age, where the predictive value of REVS and trauma on dissociation differs as a function of age (Conner, 2015; Jowett et al., 2022), potentially reflecting developmental or life stage differences in coping mechanisms and resilience. This aligns conceptually with McGuinness et al. (2025), who observed different risk-profiles across two age groups, suggesting that developmental pathways to dissociation may differ with age.

Although the effect sizes for age in the current study were small, the retention of the interaction terms provides a nuanced understanding of how age may moderate the relationship between risk factors and dissociation, emphasising that the impact of trauma and REVS is not static across lifespan. These patterns may also be influenced by the age of onset and duration of untreated dissociative symptoms (McGuinness et al., 2025), which were not accounted for in the study and should be considered in future research.

The differences between the DES-T and DES30 models illustrate how changing the outcome threshold changes the composition of cases classified as at-risk for dissociative psychopathology, and which predictors are most informative. Using the more liberal DES30 threshold nearly doubled the at-risk group (*n* = 150 [13.59%]), capturing individuals with moderate dissociative symptoms. Here, BSS-C was a significant predictor, and the BSS involuntariness perceptual factor was retained in the model. By contrast, the more conservative DES-T threshold classifies individuals with more extreme dissociative symptoms. For these severe cases, the influence of the BSS involuntariness perceptual factor emerged primarily in interaction with TEC, rather than as a standalone predictor. This plausibly indicates that, among severe cases, predictors do not operate independently but interact in predicting risk. Nonetheless, the model AUC was similar across both models, indicating a stable underlying predictive structure.

### 5.2 Limitations

Although our design allowed us to index risk for dissociative psychopathology in a large sample, the DES-T is not a rigorous substitute for clinician-administered interview schedules, which represent the gold standard for diagnosing DDs, such as the *Structured Clinical Interview for DSM Dissociative Disorders Revised* (Kundakçi et al., 2014; Steinberg, 2000). Although the current study employed a more conservative DES-T cutoff of .95, rather than the typical .90 cutoff (Waller & Ross, 1997), consideration should be given to how this threshold fares against psychiatric diagnostic assessments. In one study, the DES-T reliably identified individuals diagnosed with dissociative identity disorder via the *Dissociative Disorders Interview Schedule* (Ross et al., 1989), with a false negative rate of 5.4% (Ross, 2021). However, other research showed no relationship between taxonomic membership and clinical diagnosis, undermining the validity of the taxon as a marker of dissociative psychopathology (Modestin & Erni, 2004). Accordingly, the taxon is best interpreted as a probabilistic measure of risk for dissociative psychopathology rather than a diagnostic measure of DDs; in turn, our results should not be used for inferences regarding probable diagnoses of the at-risk group. However, we also replicated the results using an alternative cut-off, based on previous research with DES total scores (e.g., Lyssenko et al., 2018) and thus any limitations of the present work are arguably specific to the use of a self-report measure rather than the DES-T *per se*. Moreover, the implications of the model developed in the current study should be interpreted considering the absence of external validation, and that the model was tested and trained in a non-clinical sample.

### 5.3 Clinical Implications and Future Directions

Our findings indicate that incorporating the assessment of REVS into a broader battery of measures of trauma and demographic variables, may facilitate the early identification of individuals at risk for a DD. Such an approach could support timely implementation of early, targeted interventions and promote positive clinical outcome trajectories of such individuals (Shakya et al., 2024), including reduced comorbidities and suicidal tendencies commonly associated with dissociative psychopathology.

Nevertheless, the present work warrants independent replication and extension through the inclusion of more refined measures of childhood trauma, and indices of relevant mental health symptoms (McGuinness et al., 2025). Corroboration of these results via psychiatric interviews (Steinberg, 1993) would provide further information as to the predictive value of the retained variables, namely whether they are better predictors of risk probability or diagnoses. Considering the limited evidence currently available for assessing the risk of dissociative psychopathology using these factors, the data-driven approach adopted may support advances in practice. However, routine implementation of such models within clinical practice is yet to come to fruition due to an absence of sufficient external validation (Meehan et al., 2020).

### 5.4 Conclusion

The current study has developed a multivariate model that predicts the risk for dissociative psychopathology through the incorporation of trauma, REVS, and demographic measures. The findings broadly corroborate the extant literature and reinforce the importance of elevated REVS, particularly involuntariness during response to suggestion, as a risk factor for severe dissociation. With independent replication and extension, the model may help to lay the groundwork for improved early detection of individuals at risk for dissociative psychopathology with broad clinical implications spanning diagnosis and early intervention.

## Data Availability

The data is publicly available

https://osf.io/sa6bw

## Acknowledgements

This work was supported by funding from the Bial Foundation (70/16) and the Gyllenbergs Foundation to DBT.

## Open access

For the purposes of open access, the author has applied a Creative Commons Attribution (CC BY) license to any Accepted Author manuscript version arising from this submission.

## Contributors

All authors conceived the project. MVS and LW collected the data. RM analysed the data. RM drafted the initial manuscript. All authors reviewed and approved the final version of the manuscript.

