## Supplementary File for "Developing a prediction model for the risk of dissociative psychopathology from trauma and trait responsiveness to verbal suggestion"

Rebecca Morris ^a^

Madeline V. Stein ^b^

Lillian Wieder ^b^

Devin B. Terhune ^b^

^a^ *Centre for Research in Eating and Weight Disorders (CREW),* *Department of Psychological Medicine, Institute of Psychiatry, Psychology & Neuroscience, King’s College London*

^b^ *Department of Psychology, Institute of Psychiatry, Psychology & Neuroscience, King’s College London*

**Correspondence address:**

Devin B. Terhune

Department of Psychology

Institute of Psychiatry, Psychology & Neuroscience (IOPPN)

King’s College London

16 De Crespigny Park

London, UK SE5 8AB

**Open access**: For the purposes of open access, the author has applied a Creative Commons Attribution (CC BY) license to any Accepted Author manuscript version arising from this submission.

**Factor Analysis**

Bifactor and correlated factor analyses were conducted on the BSS behavioural and BSS involuntariness items to derive factor scores for inclusion in the prediction model. Model fit indices can be seen in Table S1. During the factor analysis on BSS behavioural subscale, item three was removed due to convergence error issues. Based on model fit indices and parsimony, the one-factor solution was identified as the best fit for BSS behavioural scores, with all five items loading onto the general factor (BSS behavioural general factor). By contrast, the analyses on the BSS involuntariness item scores indicated that a two-factor solution provided the best fit. As can be seen in Table S2, all items loaded onto a general factor (BSS involuntariness general factor) and two items (dreams and auditory hallucination) also loaded onto a second subordinate factor (BSS involuntariness perceptual factor).

**Table S1**

*Summary of model fit indices of bifactor and correlated factor models for BSS behavioural and involuntariness subscales (N=1,104).*

| Model | $\chi^{2}$(*p*) | | | CFI | TLI | | | | RMSEA | | | SRMR | | BIC |
| --- | --- | --- | --- | --- | --- | --- | --- | --- | --- | --- | --- | --- | --- | --- |
| BSS: behavioural | | | | |  | | |  | |  | | |  | |
| **One-factor bifactor model** | **26.948 (0.001)** | | | **0.979** | **0.958** | | | | **0.063** | | | **0.029** | | **560.503** |
| Two-factor bifactor model | Model did not converge | | | | | | | | | | | | | |
| One-factor exploratory correlated factors model | 24.779 (0.0002) | | | 0.977 | 0.954 | | | | 0.060 | | | 0.029 | | 560.503 |
| BSS: involuntariness | | | | |  | | | |  | | |  | |  |
| One-factor bifactor model | 291.400 (0.00) | | | 0.848 | 0.746 | | | | 0.169 | | | 0.072 | | 23480.768 |
| **Two-factor bifactor model** | **45.981 (0.00)** | | | **0.977** | **0.915** | | | | **0.098** | | | **0.02** | | **23360.503** |
| One-factor exploratory correlated model | 262.879 (0.00) | | | 0.831 | 0.719 | | | | 0.160 | | | 0.072 | | 23570.888 |
| Two-factor exploratory correlated factors model | | 49.934 (0.00) | 0.969 | | | 0.886 | 0.102 | | | | 0.02 | | 23360.503 | |

*Notes*. CFI = comparative fit index, TLI = tucker-lewis index, RMSEA = root mean square error of approximation, SRMR = standardised root mean squared residual, BIC= Bayesian information criterion. The best fitting model is in bold.

**Table S2**

*Factor loadings for bifactor models of BSS subscales*

|  | BSS subscale | | |
| --- | --- | --- | --- |
|  | Behavioural | Involuntariness | |
| BSS item | General factor | General factor | Perceptual factor |
| 1 (arm heaviness) | **0.57** | **0.75** | -0.35 |
| 2 (dream) | **0.43** | **0.50** | **0.39** |
| 3 (moving hands together) | N/I | **0.72** | -0.27 |
| 4 (eye catalepsy) | **0.72** | **0.65** | 0.18 |
| 5 (arm paralysis) | **0.84** | **0.72** | -0.00 |
| 6 (auditory hallucination) | **0.23** | **0.34** | **0.46** |

*Note*. Bolded values reflect the factor upon which the respective item loads. BSS = Behavioural Suggestibility Scale; N/I = Not Included

**Prediction modelling using the DES30 model**

We repeated the elastic net regression model using DES>30 as our cut-off for risk for dissociative psychopathology in order to circumvent potential limitations of the DES-T. Assumptions of elastic net logistic regression were met, with a sample size of 1104, independent observations, no multicollinearity, and a binary outcome variable. Using the DES>30 criterion, the number of people classified as at-risk for dissociative psychopathology nearly doubled (*n* = 150 [13.59%]). Elastic net logistic regression with 10-fold cross-validation identified the optimal alpha (α = 0.5) and lambda (λ = 0.006), retaining nine of the ten predictors (see Table S3). Eight retained predictors were the same as the DES--T model (Age, BSS-C, TEC, BSS behavioural general factor, BSS involuntariness perceptual factor, BSS-C x TEC, Age x TEC, Age x BSS-C); the final retained predictor was the BSS involuntariness general factor, which was not retained in the DES-T model. Thus, the DES30 model did not retain the BSS involuntariness perceptual factor x TEC interaction as seen in the DES-T model. The ROC curve yielded an AUC of 0.76 [95% CI: .72, .80], indicating a 76% probability that the model correctly classifies individuals regarding their risk for dissociative psychopathology (see Figure S1). Although nine variables were retained, BSS-C and age were the only significant predictors. Regarding overall model performance, the brier score of 0.1 reflects excellent model fit. The optimal cutoff, determined by the Youden Index (*J* = .40), indicates good discriminatory ability of the model, with good sensitivity (65%) but modest specificity (76%).

**Table S3.**

*Elastic Net Logistic Regression Model Coefficients and Odds Ratios with 95% CI for the retained predictor variables in the prediction of risk for dissociative psychopathology (N = 1,104).*

| **Variable** | **Coefficient (β)** | **OR** | **95% CI (OR)** | |
| --- | --- | --- | --- | --- |
|  |  |  | **Lower** | **Upper** |
| (Intercept) |  |  |  |  |
| Age | -0.06 | 0.95 | 0.92 | 0.96 |
| BSS-C | 0.49 | 1.64 | 1.10 | 2.31 |
| TEC | 0.29 | 1.33 | 1.00 | 1.69 |
| BSS behavioural general factor  BSS involuntariness general factor | 0.17  -0.19 | 1.18  0.83 | 1.00  0.64 | 1.53  1.00 |
| BSS involuntariness perceptual factor | 0.22 | 1.24 | 1.00 | 1.54 |
| BSS-C x TEC | -0.07 | 0.94 | 0.77 | 1.07 |
| Age x TEC | 0.004 | 1.00 | 1.00 | 1.01 |
| Age x BSS-C | 0.004 | 1.00 | 1.00 | 1.02 |

*Notes.* BSS = Behavioural Suggestibility Scale; BSS-C = Behavioural Suggestibility Scale Composite; TEC = Trauma Experiences Scale

**Figure S1**

*Receiver operating characteristic (ROC) curve of the prediction model of risk for dissociative psychopathology (N = 1,104).*

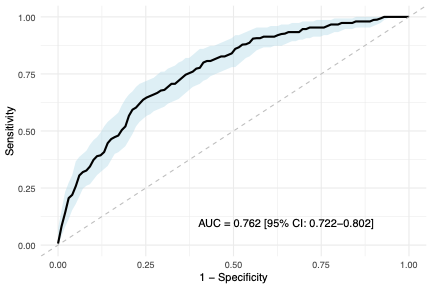

*Note.* Receiver operating characteristic (ROC) curve for the prediction model plots the true positive rate (sensitivity) against the false positive rate (1- specificity). The solid diagonal line represents random chance level discrimination.
